# Misdiagnosis prevents accurate monitoring of transmission and burden for sub-critical pathogens: a case study of *Plasmodium knowlesi* malaria

**DOI:** 10.1101/2021.09.13.21263501

**Authors:** John H. Huber

## Abstract

Maintaining surveillance of emerging infectious diseases presents challenges for monitoring their transmission and burden. Incomplete observation of infections and imperfect diagnosis reduce the observed sizes of transmission chains relative to their true sizes. Previous studies have examined the effect of incomplete observation on estimates of pathogen transmission and burden. However, each study assumed that, if observed, each infection was correctly diagnosed. Here, I leveraged principles from branching process theory to examine how misdiagnosis could contribute to bias in estimates of transmission and burden for emerging infectious diseases. Using the zoonotic *Plasmodium knowlesi* malaria as a case study, I found that, even when assuming complete observation of infections, the number of misdiagnosed cases within a transmission chain for every correctly diagnosed case could range from 0 (0 – 4) when *R*_0_ was 0.1 to 86 (0 – 837) when *R*_0_ was 0.9. Data on transmission chain sizes obtained using an imperfect diagnostic could consistently lead to underestimates of *R*_0_, the basic reproduction number, and simulations revealed that such data on up to 1,000 observed transmission chains was not powered to detect changes in transmission. My results demonstrate that misdiagnosis may hinder effective monitoring of emerging infectious diseases and that sensitivity of diagnostics should be considered in evaluations of surveillance systems.

## INTRODUCTION

For pathogens with sub-critical transmission (i.e., *R*_0_ < 1), a robust surveillance system that identifies and correctly diagnoses infections is necessary to monitor changes in pathogen transmission and burden (1). Such pathogen surveillance is important both for measuring progress towards elimination of diseases with immediate public health importance, such as measles (2–4) and malaria (5), and for assessing the future threat of emerging infectious diseases (6), such as avian influenza (7), human monkeypox (1,8), and Middle East respiratory syndrome coronavirus (2,9).

Considerable work has been devoted to advance a mathematical framework that leverages the data collected by surveillance systems to obtain estimates of transmission and burden for pathogens with sub-critical dynamics (1,2,4,10,11). These studies have improved our understanding of a wide range of emerging infectious diseases and have critically evaluated the sensitivity of these estimates to the quality of data from the surveillance system. Crucially, each study modeled variation in surveillance quality through variation in the ascertainment fraction (i.e., the proportion of infections that are detected) and assumed that, once detected, all infections were correctly diagnosed. In reality, however, non-specific clinical and biological features are likely to limit the sensitivity of clinical diagnosis, particularly for emerging infectious diseases (12,13). The extent to which misdiagnosis affects estimates of transmission and burden for pathogens with sub-critical dynamics remains largely unaddressed.

The zoonotic *Plasmodium knowlesi* malaria offers a natural case study to examine the impact of misdiagnosis on estimates of transmission and burden. Endemic to Southeast Asia (14), *P. knowlesi* is a vector-borne disease with most or all infections in humans caused by spillover transmission from the long- and pig-tailed macaque reservoir (15,16). The extent of transmission between humans is currently unknown (17). Due to morphological similarities with other *Plasmodium* spp., *P. knowlesi* is routinely misdiagnosed by light microscopy (18). A recent systematic review and meta-analysis estimated that the sensitivity of light microscopy for diagnosing *P. knowlesi* infections was less than 1% (19). This high rate of misdiagnosis greatly affects the quality of surveillance data on *P. knowlesi*, potentially biasing estimates of transmission and burden.

In this study, I aimed to evaluate the extent to which misdiagnosis of a pathogen affected the ability to monitor its change in transmission and burden. Using *P. knowlesi* as a case study, I leveraged an established framework based upon branching process theory to first quantify the potential magnitude of underestimation of pathogen burden on account of misdiagnosis. Next, I considered how underestimates of pathogen burden could lead to bias in estimates of *R*_0_, the basic reproduction number. Finally, I quantified the degree to which misdiagnosis reduced the statistical power to detect changes in transmission from surveillance data for emerging infectious diseases, such as *P. knowlesi*.

## METHODS

### Branching Process Framework of Sub-Critical Transmission

To explore the effects of misdiagnosis on the monitoring of sub-critical transmission (i.e., *R*_0_ < 1) of *P. knowlesi*, I extended a framework that uses branching process theory to estimate a pathogen’s *R*_0_ from its size distribution of stuttering transmission chains. Here, I followed Blumberg and Lloyd-Smith (1,11) and defined a transmission chain as a primary infection (i.e., a spillover infection from a zoonotic reservoir) and all secondary infections arising from that primary infection through at least one generation of pathogen transmission.

Assuming that the number of secondary infections caused through one generation of pathogen transmission followed a negative binomial distribution with mean *R*_0_ and dispersion parameter *κ*, I used the branching framework to calculate summary statistics of the transmission chains. Specifically, I solved for the probability that a transmission chain was truly of size *j* infections, *r*_*j*_, and the mean size of transmission chains, *μ*.

Following Blumberg and Lloyd-Smith (11), I considered two models of observation of infections: (i) independent observation and (ii) size-dependent observation. The model of independent observation assumes that each infection is subject to an independent probability of observation and correct diagnosis, *p*_*ind*_, that is equal to the product of the observation probability, *p*_*det*_, and the diagnostic sensitivity, *se*. By comparison, the model of size-dependent observation assumes that observation of transmission chains occurs through sentinel infections. Each infection within a transmission chain is a sentinel infection with probability, *p*_*sent*_, and, if there is at least sentinel infection within the transmission chain, then all infections within the transmission chain are observed. Diagnosis of each infection occurs independently and is subject to sensitivity, *se*.

I then computed the mean observed transmission chain size, *μ*^∗^, as a function of the transmission parameters (*R*_0_ and *κ*), the observation model (*p*_*det*_ or *p*_*sent*_), and the diagnostic accuracy (*se*). This allowed me to relate the distribution of observed transmission chain sizes to the distribution of true transmission chain sizes and quantify bias in the maximum-likelihood estimates of transmission, 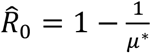. If all infections are observed and correctly diagnosed, then *μ*^∗^ = *μ* and thus 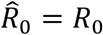. Violations of this assumption, either through incomplete observation or misdiagnosis, introduce bias into transmission estimates. A full description of the branching process framework can be found in the Supplement.

### Analyses

#### Quantifying the Bounds of Total Burden

To first demonstrate how misdiagnosis, in addition to incomplete observation, may lead to an underestimate of *P. knowlesi* burden, I computed the probability distribution of the true size of a transmission chain conditional upon the observed size of a transmission chain. That is, given that I observed a transmission chain of size *ĵ*, the probability that the transmission chain is truly of size *j* is equal to

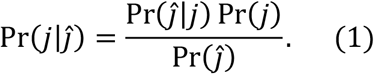

In eq. (1), Pr (*j*) is the probability that a transmission chain is of size *j, r*_*j*_, computed using eq. (S1), and Pr (*ĵ*) is the probability that a transmission chain is of observed size *j*, 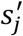, computed using eq. (S2) for the model of independent observation and using the numerator of eq. (S7) for the model of size-dependent observation. For the model of independent observation, the probability of observing a transmission chain of size *ĵ* given that the transmission chain is truly of size *j* is

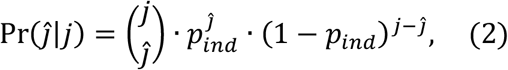

where *p*_*ind*_ is equal to the product of the probability of detection, *p*_*det*_, and the sensitivity of diagnosis, *se*. By comparison, for the model of size-dependent observation, this quantity is computed as

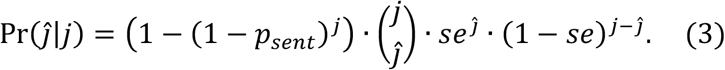

Substituting the respective terms into eq. (1), for the model of independent observation, I computed the probability that a transmission chain is of true size *j* given that it is observed to be of size *ĵ* as

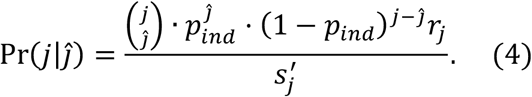

For the model of size-dependent observation, I computed this quantity as

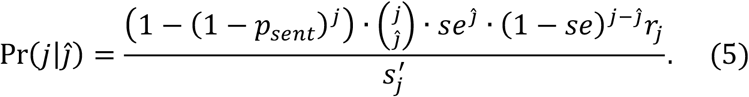

I used eqs. (4-5) to compute the expected true transmission chain sizes given observed transmission chains of one, two, or three cases while varying the probability of observation, *p*_*det*_ or *p*_*sent*_, from 0.1 to 1.0 in increments of 0.1. I sampled the sensitivity of the diagnostic method from the posterior estimate of sensitivity of light microscopy for *P. knowlesi* with mean equal to 1.19 × 10^−3^ (19). For each combination of observed chain size and probability of observation, I calculated the expected true transmission chain size, assuming that the true value of *R*_0_ was equal to 0.1, 0.5, or 0.9. These values of *R*_0_ represent low, medium, and high values of sub-critical transmission and fall within the plausible range of human-to-human transmission of *P. knowlesi* (20). The dispersion parameter *κ* was assumed to be 0.1 in all scenarios, though a supplementary analysis was performed where *κ* → ∞.

#### Effect of Misdiagnosis on Estimates of Transmission

On account of incomplete observation and misdiagnosis, the observed burden of *P. knowlesi* may not reflect the true burden. It follows that the mean observed transmission chain size will not equal the true mean transmission chain size, biasing our estimates of *R*_0_. To explore the extent of this bias in scenarios where *R*_0_ was equal to 0.1, 0.5, or 0.9, I calculated the mean observed transmission chain sizes while varying the probabilities of observation, *p*_*det*_ or *p*_*sent*_, from 0.1 to 1.0 in increments of 0.1 and while using posterior samples of sensitivity of light microscopy for *P. knowlesi* (19). I then compared the maximum-likelihood estimates of 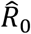 to the true values of *R*_0_ for both the models of independent observation and size-dependent observation. In all scenarios, I assumed that the dispersion parameter *κ* was 0.1, and a supplementary analysis was performed were *κ* → ∞.

#### Effect of Misdiagnosis on Statistical Power to Detect Changes in Transmission

Bias in *R*_0_ estimates on account of misdiagnosis could reduce the statistical power to detect changes in *R*_0_ over time using data on the size of transmission chains. To measure statistical power as a function of the number of observed transmission chains, I followed an approach taken by Blumberg *et al*. (2). I assumed that *R*_0_ was historically equal to 0.1 and then increased to *R*_0_ + Δ*R*_0_, where Δ*R*_0_ was set to 0.1, 0.5, or 0.9. I then simulated *N* observed transmission chains and estimated 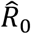 while varying *N* from 1 to 1,000. I then compared the model in which *R*_0_ was estimated to have changed to the null hypothesis that there was no change in transmission (i.e., Δ*R*_0_ = 0) using the Akaike Information Criterion (AIC) (21). For each number of observed transmission chains *N*, I repeated this procedure 1,000 times and computed statistical power as the proportion of simulations in which I detected a change in transmission on the basis of AIC. To measure the minimum effect of misdiagnosis on statistical power, I set *p*_*det*_ and *p*_*sent*_ equal to 1. Because all infections are observed under this assumption and diagnosis is performed independently across infections in both models, the models of independent and size-dependent observation yield identical results. In all scenarios, I assumed that the dispersion parameter *κ* was 0.1, and a supplementary analysis was performed were *κ* → ∞.

## RESULTS

Assuming complete observation of infections (i.e., *p*_*det*_ and *p*_*sent*_ equal to 1), misdiagnosis of *P. knowlesi* infections would underestimate the true *P. knowlesi* burden, with the magnitude of this effect depending upon *R*_0_ (Fig. 1). For a scenario in which *R*_0_ was 0.1, the expected true size of a transmission chain is one infection (95% CI: 1 – 5) if the observed size is one case, four infections (2 – 12) if the observed size is two cases, and seven infections (3 – 17) if the observed size is three cases. Under an alternative scenario in which *R*_0_ was 0.9, the expected true size of the transmission chains increased to 87 (1 – 838), 461 (49 – 965), and 650 (157 – 983) infections, respectively.

**Figure 1.**
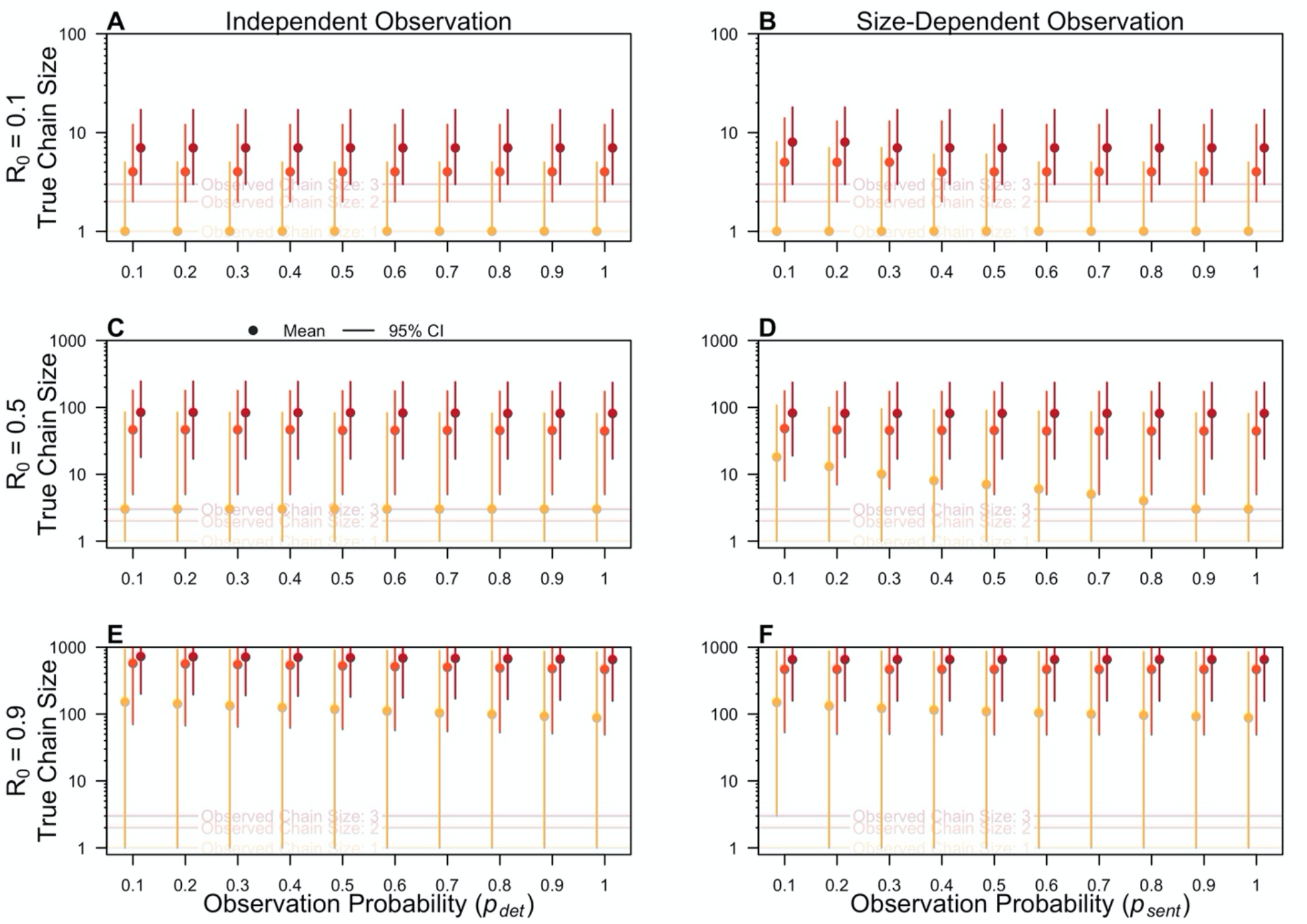
Effect of misdiagnosis and imperfect observation on the expected burden. The mean true transmission chain size (dots) and 95% CI (segments) are shown conditional upon on an observed transmission chain size of one (yellow), two (orange), or three (red) cases and an R_0_ of 0.1 (A,B), 0.5 (C,D), and 0.9 (E,F). The horizontal axis is the observation probability, representing p_det_ for the Model of Independent Observation (A, C, E) and p_sent_ for the Model of Size-Dependent Observation (B, D, F).

The effect of incomplete observation (i.e., *p*_*det*_ or *p*_*sent*_ < 1) on the expected burden was most apparent at an intermediate *R*_0_ of 0.5 and with the model of size-dependent observation (Fig. 1D). Under this scenario, given a transmission chain of size one, the expected true transmission chain was 18 infections (1 – 107) if *p*_*sent*_ was equal to 0.1, compared to 3 infections (1 – 80) if *p*_*sent*_ was equal to 1. In all other scenarios, the expected burden did not change significantly with *p*_*det*_ or *p*_*sent*_. This occurred because, even with complete observation (i.e., *p*_*det*_ and *p*_*sent*_ equal to 1), 99.881% of *P. knowlesi* cases were expected to be misdiagnosed, given a sensitivity of 0.119%. Therefore, irrespective of the observation probability, only a subset of true transmission chain sizes is consistent with the sizes of the observed transmission chains, given this high percentage of false negatives. If I instead assumed perfect sensitivity of the method, I observed a greater effect of the observation probability on the expected burden (Fig. S1).

Given that misdiagnosis underestimated the burden of *P. knowlesi* in this analysis, I assessed its effect on my estimates of transmission, 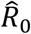. Because misdiagnosis caused the average observed size of transmission chains to be less than the average true size of transmission chains, I consistently underestimated 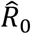, with the effect being more severe at lower *R*_0_ (Fig. 2). For example, with an *R*_0_ of 0.1 and assuming perfect observation of infections (i.e., *p*_*det*_ and *p*_*sent*_ equal to 1), my median 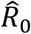 estimate was 1.9 × 10^−4^ (95% PPI: 2.0 × 10^−5^ – 2.9 × 10^−3^), corresponding to a 520-fold (34 – 4900) underestimate of transmission. Under an alternative scenario in which *R*_0_ was 0.9, my median 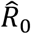 was 0.26 (0.046 – 0.66), corresponding to a 3.5-fold (1.4 – 19.4) underestimate in transmission.

**Figure 2.**
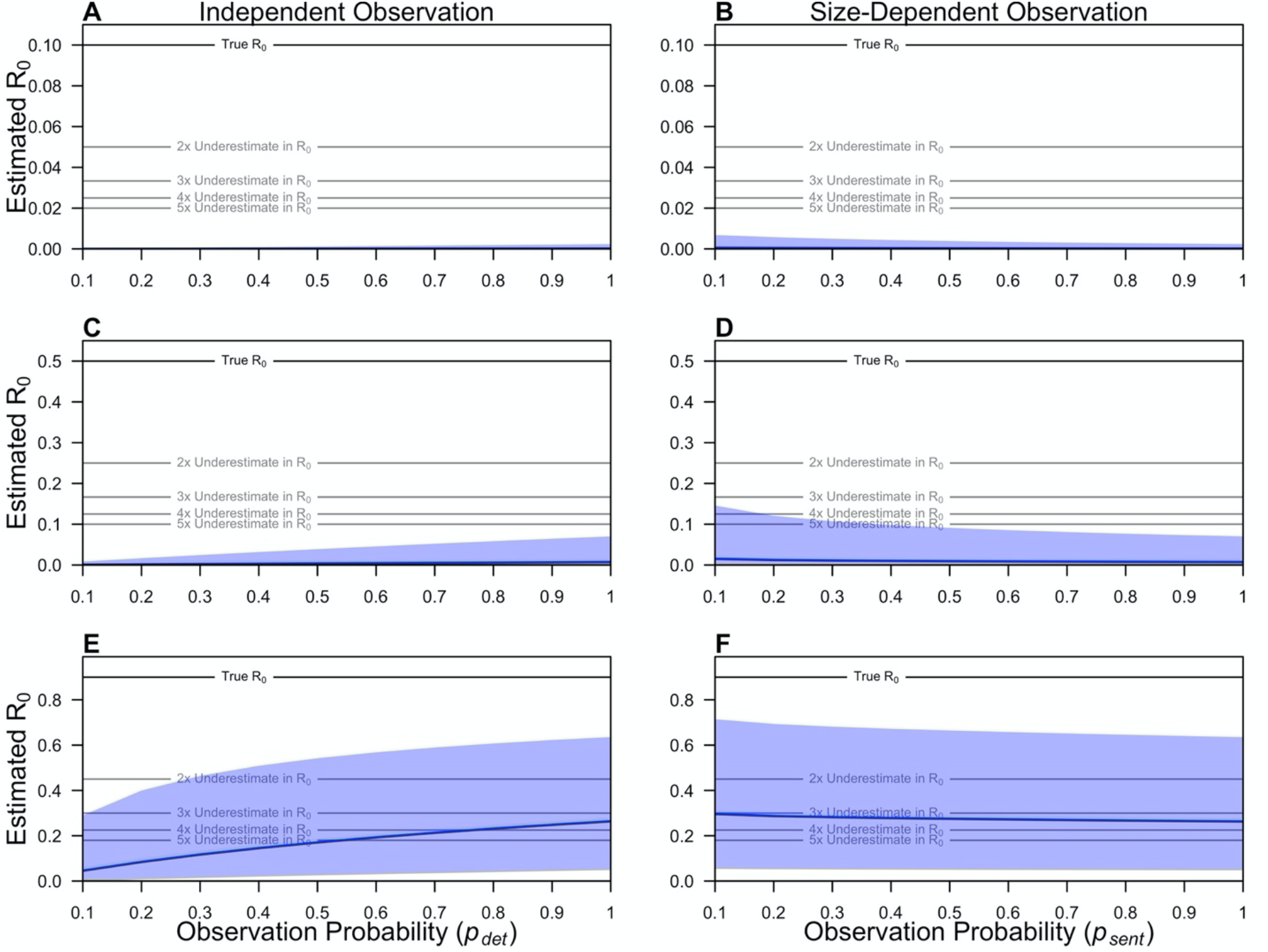
Effect of misdiagnosis and imperfect observation on estimates of transmission. The posterior median (blue line) and 95% posterior prediction interval (blue shaded region) of maximum-likelihood estimates of R_0_ are shown as a function of the observation probability. The observation probability represents p_det_ for the Model of Independent Observation (A, C, E) and p_sent_ for the Model of Size-Dependent Observation (B, D, F). The solid black denotes the true R_0_ in each panel, and the grey lines denote two-to-five-fold underestimates of R_0_ in each panel.

My estimates of transmission were sensitive to the simulated observation probability, though the direction of the effect depended upon the assumed model of observation. For the model of independent observation, 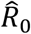 estimates increased with increasing *p*_*det*_, because the average observed size of transmission chains increased as more infections were observed (Fig. 2, left column). By contrast, 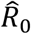 estimates decreased with increasing *p*_*sent*_ for the model of size-dependent observation (Fig. 2, right column). This counterintuitive effect can be explained by the observation that, if *p*_*sent*_ is low, larger transmission chains have a greater probability that at least one infection is a sentinel infection. This causes a bias in the size of the transmission chains that are observed at low values of *p*_*sent*_, increasing the mean observed transmission chain size relative to that at higher values of *p*_*sent*_ and thus inflating the 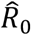 estimate.

The underestimates of burden (Fig. 1) and transmission (Fig. 2) indicated that misdiagnosis of *P. knowlesi* may affect the statistical power to detect changes in transmission based on the size of observed transmission chains. To test this, I simulated changes in transmission and measured the statistical power to detect that change. I observed that, under scenarios in which *R*_0_ increased by 0.9, data on 1,000 observed transmission chains provided only 10.3% power using an imperfect diagnostic method, compared to 100% if using a perfect diagnostic method (Fig. 3). At smaller increases in *R*_0_, data on observed transmission chain sizes obtained using an imperfect diagnostic method had effectively no power to detect a change in transmission.

**Figure 3.**
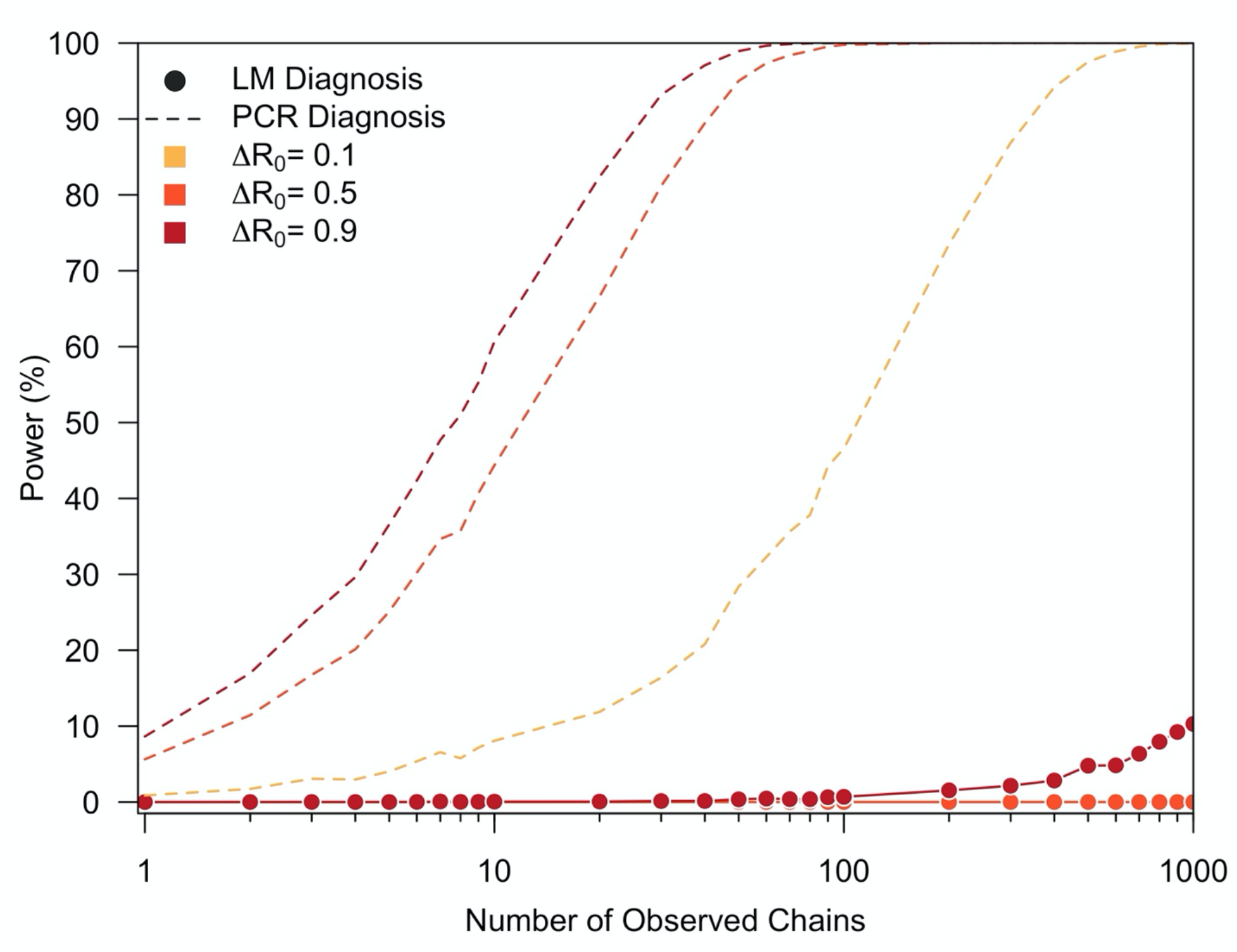
Effect of misdiagnosis on the statistical power to detect changes in transmission. The statistical power (%) to detect an increase in transmission is shown as a function of the number of observed chains for a transmission increase (ΔR_0_) of 0.1 (yellow), 0.5 (orange), and 0.9 (red). Solid lines and points represent an imperfect diagnostic method (i.e., LM) and the dotted lines represent perfect diagnosis (i.e., PCR).

## DISCUSSION

Obtaining accurate estimates of transmission and burden is important for monitoring the emergence of infectious diseases. Previous studies have explored the extent to which incomplete observation of infections affects the estimates of transmission for such pathogens (1,2,7,11). In this work, I built upon these studies by considering the effect of misdiagnosis on estimates of pathogen transmission and burden. Using the zoonotic *P. knowlesi* malaria as a case study, I found that misdiagnosis—independent of incomplete observation of infections—may cause us to underestimate the transmission and burden of pathogens with sub-critical dynamics and hinders effective, prospective monitoring of changes in transmission.

My results demonstrate that, for pathogens with sub-critical transmission, misdiagnosis leads to an underestimate of overall pathogen burden. Depending upon the *R*_0_ simulated, I found that there could be as many as 86 misdiagnosed cases, on average, for each correctly diagnosed case of *P. knowlesi*, even when assuming complete observation of infections. This effect increased under select settings when the assumption of complete observation of infections was relaxed. The underestimation of burden due to misdiagnosis has the potential to shape our epidemiological understanding of an emerging pathogen. For instance, singleton cases of a zoonotic pathogen, such as *P. knowlesi*, are commonly assessed as dead-end spillover events from the zoonotic reservoir (17). However, my simulations suggest that such singleton cases could instead represent a broad range of epidemiological outcomes, spanning dead-end spillover events to larger transmission chains.

Due to its effect on observed pathogen burden, misdiagnosis contributed a downward bias in estimates of transmission. Except for scenarios in which the true simulated *R*_0_ was close to one, my maximum-likelihood estimates of *R*_0_ approached zero, representing situations in which we would incorrectly conclude that human-to-human transmission of the pathogen was unlikely to be occurring. For every scenario considered, the estimate of *R*_0_ was less than the true value, indicating that bias due to misdiagnosis exceeds the competing positive bias from incomplete observation when assuming size-dependent observation (1). For pathogens such as *P. knowlesi*, these simulation results suggest that, in settings where misdiagnosis is common, the extent of human-to-human transmission could be greater than previously thought. To date, it has been believed that nearly all cases of *P. knowlesi* in humans are caused by spillover from long-tailed and pig-tailed macaques, the zoonotic reservoir (17). The lack of observed human-to-human transmission may be explained by multiple factors, including low parasite densities in humans (16) and restricted vector habitat preference (15), and is supported by a lack of genetic diversity across human *P. knowlesi* infections (22). Nevertheless, human-to-human transmission of *P. knowlesi* has been demonstrated experimentally (23), and these results suggest that, if or when human-to-human transmission occurs, misdiagnosis could cause us to underestimate its magnitude.

Finally, I demonstrated that data on the sizes of transmission chains diagnosed using a diagnostic with realistic sensitivity would be insufficient to monitor changes in transmission. Even with 1,000 observed transmission chains, I calculated a power of only 10% to detect an increase in *R*_0_ from 0.1 to 1. This empirical power calculation assumed complete observation of infections, so it represents an upper bound on the statistical power that we might expect if a diagnostic with realistic sensitivity was used. Therefore, more sensitive diagnostics, such as polymerase chain reaction, may be needed to detect changes in transmission that could result from pathogen evolution (24), among other factors (25–27).

This analysis is subject to a number of limitations. First, the conclusions that I reached were based upon simulated data only. I used simulations representative of *P. knowlesi* to illustrate possible outcomes that may occur due to misdiagnosis (19,20), yet I lacked empirical data on the distribution of transmission chain sizes for *P. knowlesi*. As such, this analysis is not estimating the true extent of human-to-human transmission of *P. knowlesi*. Second, methods exist to account for incomplete observation of infections in estimates of *R*_0_ (11). However, as noted by Blumberg *et al*. (2), it is challenging to estimate the proportion of infections that are captured by the surveillance system. Consequently, these calculations were conditioned upon the assumption of complete observation and perfect diagnoses, so violations therein should be interpreted as the upper bound on the bias that would likely be observed. Finally, I considered a single pathogen in isolation, though misdiagnosis is commonly due to co-circulation of related pathogens (12,13,18). Accounting for the upward bias due to false-positive diagnoses from other pathogens and exploring the magnitude of this effect across epidemiological settings could be important directions for future work.

## Data Availability

The code and data to reproduce the analyses will be available upon publication.

## ACKNOWLEDGMENTS

I acknowledge support from a Graduate Research Fellowship from the National Science Foundation, a Richard and Peggy Notebaert Premier Fellowship from the University of Notre Dame, and the National Institute of General Medical Sciences (grant number 1R35GM143029-01 to Alex Perkins). The funders had no role in the study design, in the collection, analysis, and interpretation of data, in the writing of the report, or in the design to submit the article for publication. I thank Alex Perkins for helpful comments on this manuscript.

## SUPPLEMENT

### Methods

#### Mean Transmission Chain Size

##### Complete Observation and Correct Diagnosis

For a pathogen with sub-critical transmission dynamics, I modeled the number of offspring caused by a single infection through one generation of transmission as a negative binomial distribution with mean *R*_0_ and dispersion parameter *κ* (1,11). Therefore, it follows that transmission chains of size *j* occur with probability,

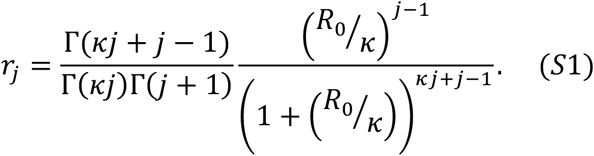

In eq. (S1), Γ(·) is the gamma function. Because *R*_0_ < 1, the mean transmission chain size *μ* can be calculated as the mean of a geometric series with common ratio *R* and is equal to 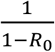.

##### Incomplete Observation and Imperfect Diagnosis

In the case of *P. knowlesi* and many other pathogens, the size of transmission chains that are identified by a surveillance system will be affected by two factors. First, infections in a transmission chain may not present within the health system, due to a lack of symptoms or access to treatment. Second, infections in the transmission chain that do present within the health system may be misdiagnosed and thus inaccurately recorded within the surveillance system. Both factors will make the observed transmission chain appear smaller in size than the true transmission chain. Previous work by Blumberg and Lloyd-Smith (1) has quantified the effect of two models of incomplete observation on estimates of transmission and burden. Here, I build upon this work by integrating the effect of (mis)diagnosis of infections that occurs secondary to the observation of infections within the health system.

###### Model of Independent Observation

The first model of incomplete observation and diagnosis assumes that each individual is subject to an independent probability *p*_*ind*_ equal to the product of observation probability, *p*_*det*_, and the sensitivity of the diagnostic method, *se*. Therefore, the probability that we observe and correctly diagnose *j* cases from a transmission chain is equal to

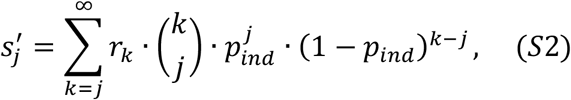

where *r*_3_ is the probability that a transmission chain is of true size *κ*, calculated using eq. (S1). The probability that a transmission chain is of observed size *j* is equal to the normalized probability of 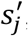, computed as

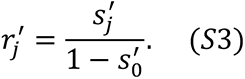

In eq. (S3), 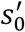 is the probability that a transmission chain is neither observed nor correctly diagnosed. Due to incomplete observation and misdiagnosis, the probability that a transmission chain is of observed size *j*, 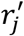, is not equal to the probability that a transmission chain is of true size *j, r*_*j*_. Finally, because each infection within the transmission chain is subject to an independent probability of observation and correct diagnosis, the probability *p*_67(_ that a randomly sampled infection is observed and correctly diagnosed is equal to *p*_*ind*_.

###### Model of Size-Dependent Observation

The alternative model assumes that transmission chains are observed through a sentinel infection, such that, if the sentinel infection is observed, then all other infections in the transmission chain will be observed. Incorporating the effect of imperfect diagnosis, the probability that we do not observe a transmission chain of size *j* is equal to

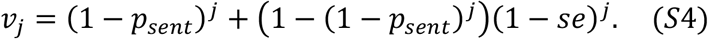

In eq. (S4), the first term in the summation is the probability that none of the *j* infections of the transmission chain are a sentinel infection, and the second term in the summation is the product of the probability that at least one infection is a sentinel infection (i.e., the probability that we observe the transmission chain) and the probability that all *j* infections are misdiagnosed. Using eq. (S4), I calculated the probability that a transmission chain is neither observed nor correctly diagnosed as

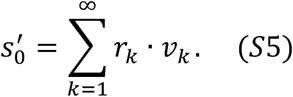

The probability that a transmission chain is of observed size *j* is then equal to

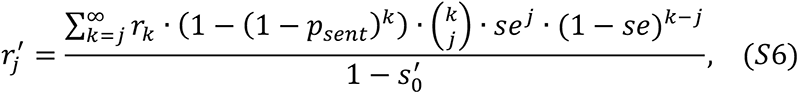

and the probability that a randomly chosen infection is observed and correctly diagnosed is equal to

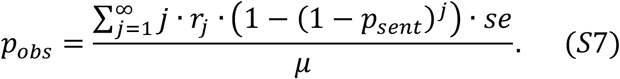

Eq. (S7) accounts for the probability that non-sentinel infections are detected, a quantity that increases as a function of the transmission chain size.

###### Mean Transmission Chain Size

For both the model of independent observation and the model of size-dependent observation, the mean observed size of transmission chains is calculated as

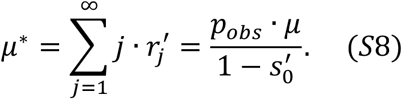

## Results

**Figure S1.**
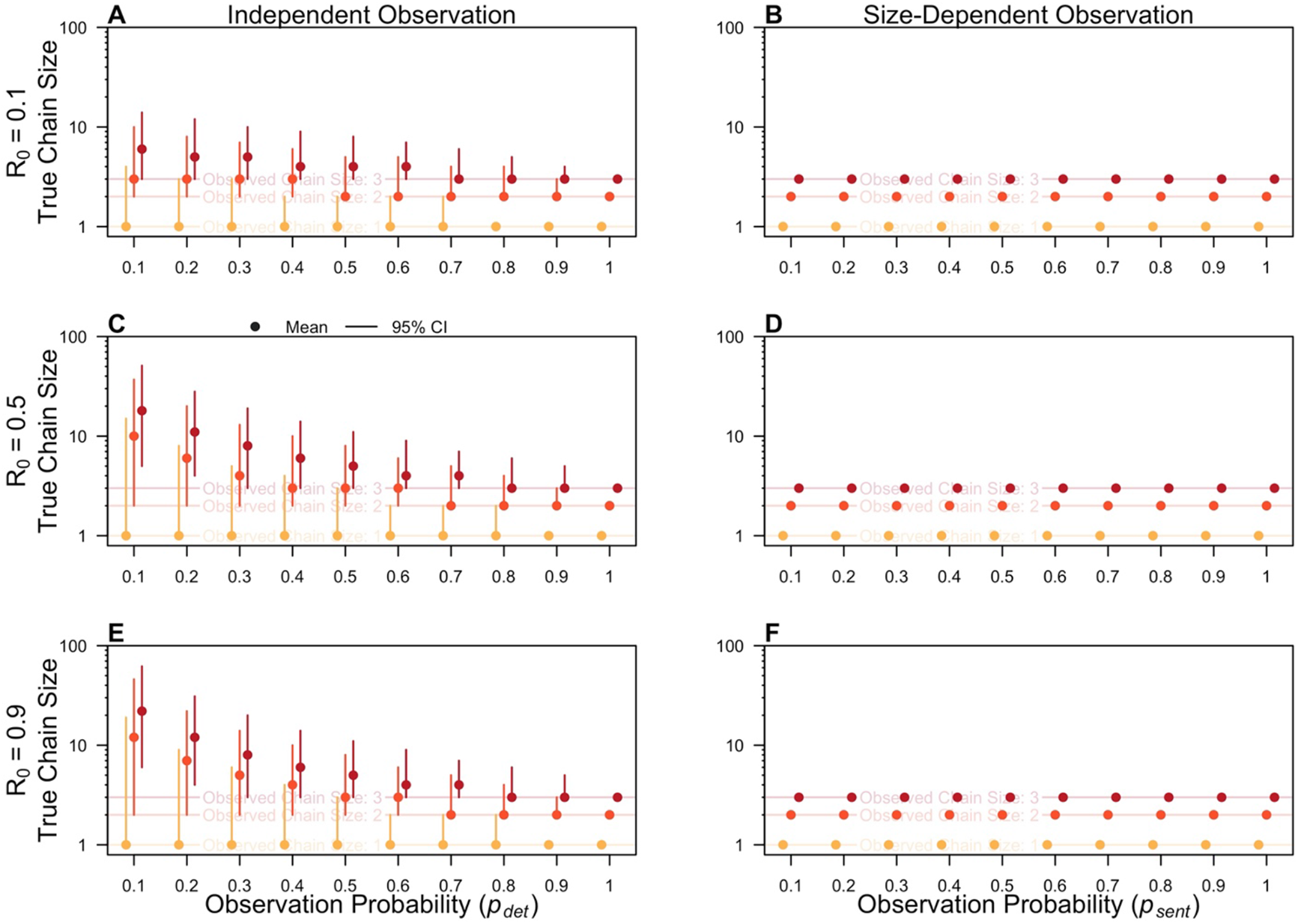
Effect of misdiagnosis and imperfect observation on the expected true pathogen burden assuming perfect diagnosis. The mean true transmission chain size (dots) and 95% CI (segments) are shown conditional upon on an observed transmission chain size of one (yellow), two (orange), or three (red) cases and an R_0_ of 0.1 (A,B), 0.5 (C,D), and 0.9 (E,F). The horizontal axis is the observation probability, representing p_det_ for the Model of Independent Observation (A, C, E) and p_sent_ for the Model of Size-Dependent Observation (B, D, F).

**Figure S2.**
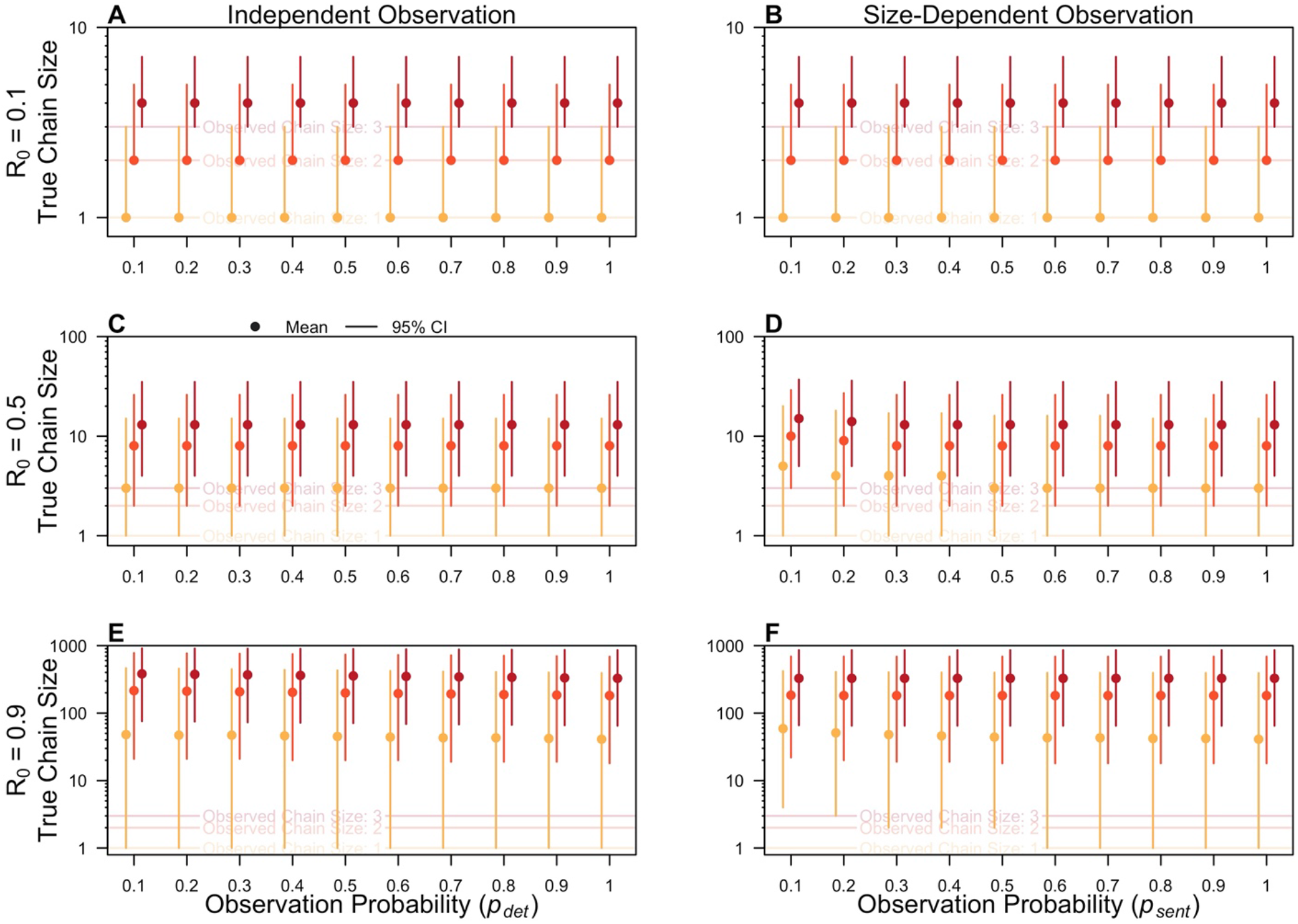
Effect of misdiagnosis and imperfect observation on the expected true pathogen burden when k κ → ∞. The mean true transmission chain size (dots) and 95% CI (segments) are shown conditional upon on an observed transmission chain size of one (yellow), two (orange), or three (red) cases and an R_0_ of 0.1 (A,B), 0.5 (C,D), and 0.9 (E,F). The horizontal axis is the observation probability, representing p_det_ for the Model of Independent Observation (A, C, E) and p_sent_ for the Model of Size-Dependent Observation (B, D, F).

**Figure S3.**
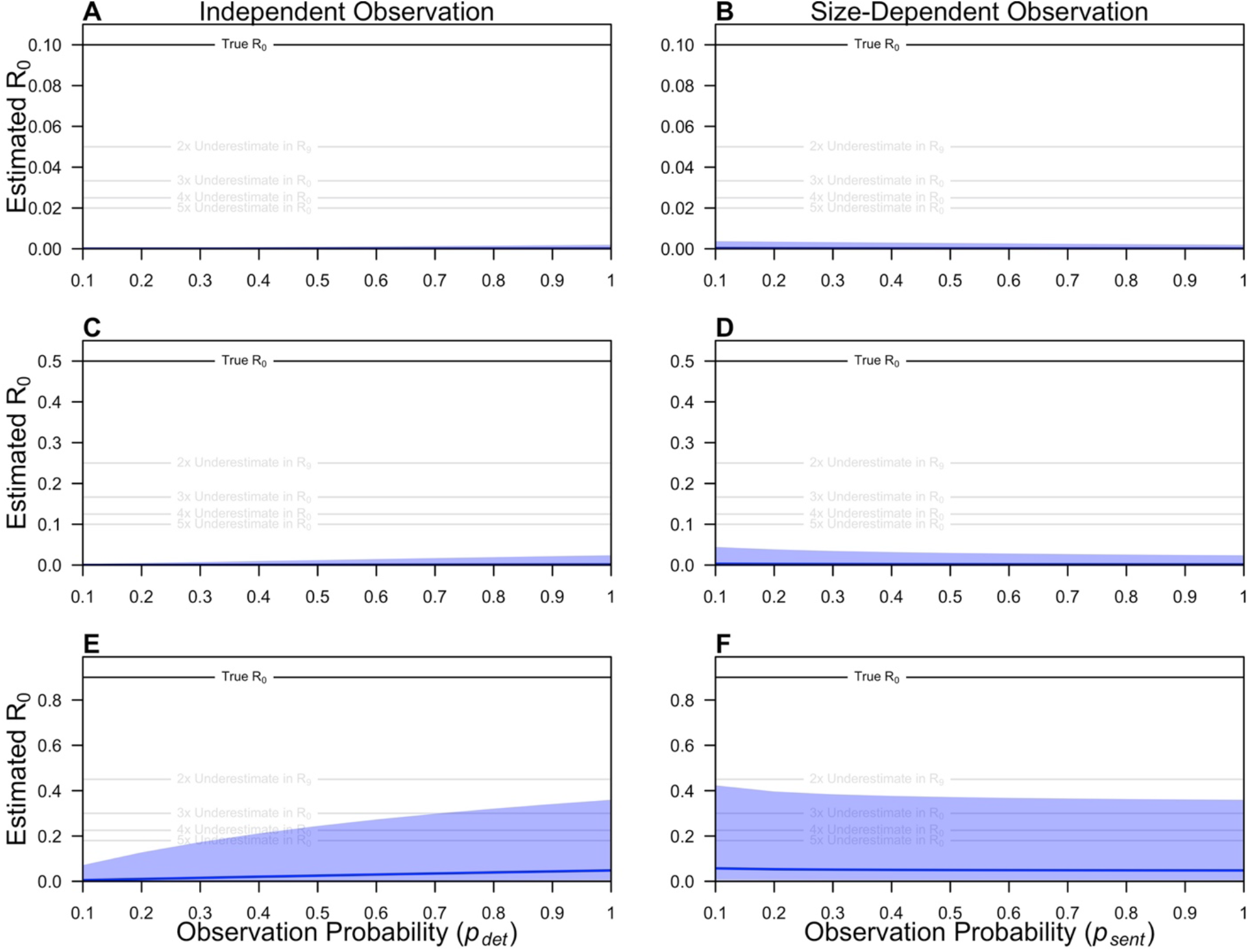
Effect of misdiagnosis and imperfect observation on estimates of transmission when k κ → ∞. The posterior median (blue line) and 95% posterior prediction interval (blue shaded region) of maximum-level estimates of R_0_ are shown as a function of the observation probability. The observation probability represents p_det_ for the Model of Independent Observation (A, C, E) and p_sent_ for the Model of Size-Dependent Observation (B, D, F). The solid black denotes the true R_0_ in each panel, and the grey lines denote two-to-five-fold underestimates of R_0_ in each panel.

**Figure S4.**
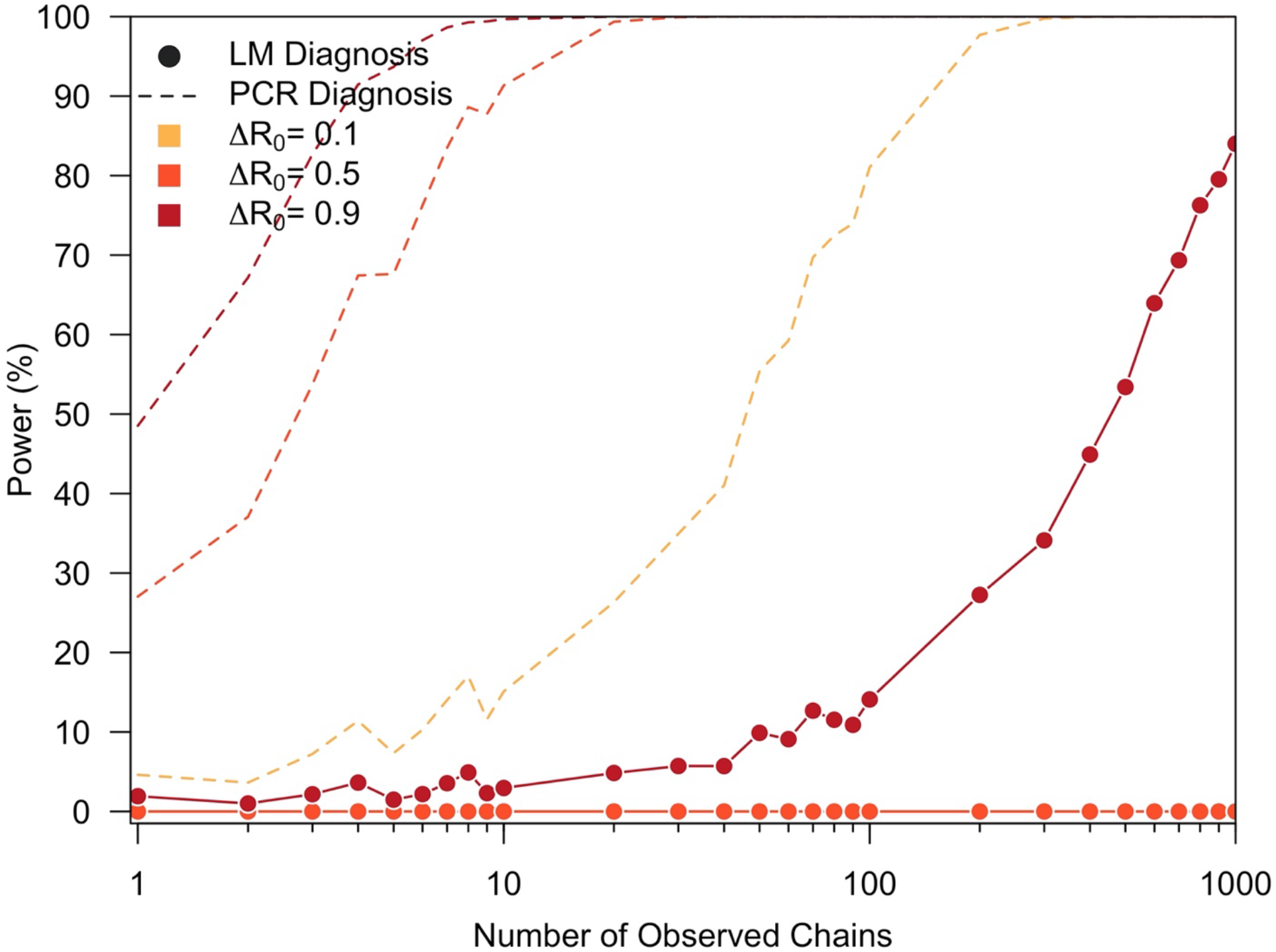
Effect of misdiagnosis on the statistical power to detect changes in transmission when k κ → ∞. The statistical power (%) to detect an increase in transmission is shown as a function of the number of observed chains for a transmission increase (ΔR_0_) of 0.1 (yellow), 0.5 (orange), and 0.9 (red). Solid lines and points represent an imperfect diagnostic method (i.e., LM) and the dotted lines represent perfect diagnosis (i.e., PCR).

